# Multi-Task TabGraphSyn: Graph-Based Synthetic EHR Generation with Improved Quality-Privacy Trade-offs for Opioid Use Disorder Prediction

**DOI:** 10.64898/2026.04.24.26351704

**Authors:** Mohammad Arif Ul Alam, Sophia Shalhout

**Affiliations:** Richard A Miner School of Computer and Information Sciences, University of Massachusetts Lowell,, Lowell,, MA, USA; Otolaryngology-Head and Neck Surgery, Harvard Medical School,, Cambridge,, MA, USA

**Keywords:** Electronic Health Records (EHR), Synthetic Data Generation, Privacy-Preserving Machine Learning, Graph Neural Networks, Diffusion Models

## Abstract

Electronic health record (EHR) data are critical for clinical research but are challenging to share due to privacy and re-identification risks, particularly in sensitive domains such as opioid use disorder (OUD). Synthetic data generation offers a promising alternative; however, existing methods often struggle to preserve complex multivariate dependencies while maintaining a strong balance between data utility and privacy. The recently proposed MIIC-SDG framework leverages multivariate information theory and Bayesian network modeling to capture dependency structures and introduces Quality-Privacy Scores (QPS) to evaluate this tradeoff, yet its capacity to model nonlinear relationships and support multi-task predictive settings remains limited. In this work, we propose a multi-task extension of TabGraphSyn, a graph-based generative framework for privacy-preserving EHR synthesis. The method constructs patient similarity graphs from high-dimensional tabular data and learns topology-aware embeddings via a graph convolutional network, which are then incorporated into a conditional variational autoencoder for synthetic data generation. Unlike prior approaches, our framework jointly models multiple clinically relevant OUD targets, including 180-day opioid abuse outcome, opioid concept group, and opioid source concept group, enabling preservation of label-dependent relationships across tasks. We evaluate TabGraphSyn against MIIC-SDG under a unified framework including multi-task predictive utility, distributional similarity, identifiability risk, membership inference risk, and QPS-based metrics. Results on the NIH All of Us dataset show that TabGraphSyn achieves a stronger overall utility-privacy balance, outperforming MIIC in most headline metrics, including higher synthetic multi-task ROC-AUC (0.5278 vs 0.4932), MetaQPS (AM: 0.0215 vs 0.0115; HM: 0.0391 vs 0.0223), while slightly underperforming in macro F1 (0.2321 vs 0.2840). These findings demonstrate improved modeling of nonlinear dependencies and more favorable quality-privacy trade-offs in multi-task settings, supporting its use for realistic and privacy-aware synthetic EHR data generation.

## 1. Introduction

The rapid digitization of healthcare systems has led to an unprecedented accumulation of Electronic Health Records (EHR), enabling data-driven clinical research, precision medicine, and large-scale epidemiological studies. However, the sensitive nature of medical data imposes strict privacy constraints, limiting data sharing and secondary usage under regulatory frameworks such as the General Data Protection Regulation (GDPR) Dwork et al. (2006). As a result, developing methods that enable safe and effective utilization of healthcare data while preserving patient privacy has become a critical challenge in modern biomedical informatics.

Traditional anonymization techniques, including k-anonymity, l-diversity, and t-closeness, attempt to mitigate re-identification risks through data generalization and suppression Sweeney (2002); Machanavajjhala et al. (2007); Li (2025). While these approaches provide formal privacy guarantees, they often distort the underlying data distribution and degrade analytical utility, especially in high-dimensional clinical datasets Hernandez et al. (2022). Moreover, such methods struggle to preserve complex multivariate dependencies, which are essential for downstream predictive modeling and clinical insight generation.

To address these limitations, synthetic data generation (SDG) has emerged as a promising alternative. SDG aims to learn the underlying data distribution and generate artificial samples that retain statistical properties of the original dataset without exposing identifiable patient information Tucker et al. (2020). Recent advances in generative modeling, including Generative Adversarial Networks (GANs) Choi et al. (2017); Baowaly et al. (2019) and Variational Autoencoders (VAEs), have demonstrated strong potential for modeling high-dimensional tabular healthcare data.

Despite these advancements, several fundamental challenges remain. First, many existing methods fail to adequately capture higher-order multivariate relationships inherent in clinical data Sella et al. (2025). Second, there exists a fundamental tradeoff between data utility and privacy: models that closely approximate real data distributions may increase the risk of reidentification or membership inference attacks Stadler et al. (2020). Third, most approaches focus on single-task settings and do not explicitly preserve label-dependent relationships across multiple clinically relevant outcomes, limiting their applicability in real-world predictive modeling scenarios.

Recent work such as MIIC-SDG leverages multivariate information theory and Bayesian network modeling to capture dependency structures and introduces Quality-Privacy Scores (QPS) to quantify the trade-off between fidelity and privacy Sella et al. (2025). While effective in modeling conditional independence relationships, such approaches are inherently limited in capturing complex nonlinear interactions and are not naturally suited for multi-task predictive settings.

Graph-based learning provides a compelling direction to address these challenges. By explicitly modeling relationships among patients or features, graph representations can capture structured dependencies beyond pairwise correlations. When combined with deep generative models, graph-based embeddings can enhance the ability to learn complex distributions while preserving relational structure.

In this work, we propose a **multi-task extension of TabGraphSyn**, a graph-based generative framework for privacy-preserving EHR synthesis. Our approach constructs patient similarity graphs from high-dimensional tabular data and learns topology-aware embeddings using a graph convolutional network (GCN). These embeddings are then integrated into a conditional variational autoencoder (VAE) to generate synthetic data that preserves both feature distributions and label-dependent relationships. Unlike prior approaches, our framework jointly models multiple clinically relevant opioid use disorder (OUD) targets, including 180-day opioid abuse outcome, opioid concept group, and opioid source concept group.

To ensure rigorous evaluation, we adopt a unified framework that assesses both utility and privacy, including multi-task predictive performance, distributional similarity, identifiability risk, membership inference risk, and Quality-Privacy Scores (QPS). Through experiments on the NIH All of Us dataset, we demonstrate that the proposed method achieves a stronger overall balance between utility and privacy compared to MIIC-SDG, highlighting its effectiveness for realistic and privacy-aware synthetic EHR data generation.

Our main contributions are summarized as follows:

- We propose a novel multi-task TabGraphSyn framework that integrates graph-based representation learning with conditional variational generation for synthetic EHR data.
- We extend synthetic data generation to a multi-task setting, enabling preservation of label-dependent relationships across multiple clinically relevant OUD outcomes.
- We provide a comprehensive evaluation aligned with the MIIC-SDG framework, incorporating multi-task utility, identifiability, membership inference, and QPS-basedmet-rics.
- We demonstrate improved utility-privacy trade-offs and stronger modeling of nonlinear dependencies compared to MIIC-SDG on a real-world clinical dataset.

## 2. Methodology

In this study, we propose a **multi-task TabGraphSyn** framework for privacy-preserving synthetic electronic health record (EHR) generation. The proposed method is designed for highdimensional clinical data, where complex dependencies exist among structured variables and clinically meaningful prediction targets. Unlike traditional anonymization methods, which often reduce downstream utility, and unlike many synthetic data generation approaches that fail to preserve multivariate structure, our framework aims to jointly support data fidelity, predictive utility, and privacy preservation Hernandez et al. (2022); Tucker et al. (2020).

Recent advances in synthetic data generation, including generative adversarial networks (GANs), variational autoencoders (VAEs), and other deep generative approaches, have shown promise for healthcare applications Choi et al. (2017); Baowaly et al. (2019). However, many of these methods treat patient records as independent tabular samples and therefore do not explicitly model inter-patient similarity structure. In contrast, graphical approaches such as MIIC-SDG preserve multivariate dependencies through information-theoretic and Bayesian network modeling, and provide a useful quality-privacy evaluation framework Sella et al. (2025). Nevertheless, such methods are less flexible in modeling complex nonlinear relationships and are not naturally formulated for multi-task predictive settings. To address these limitations, our method combines graph-aware representation learning with conditional variational generation, enabling realistic synthetic EHR generation while preserving multiple opioid-related prediction targets.

### 2.1. Overview of the Proposed Framework

Figure 1 illustrates the overall architecture of the proposed framework. The pipeline consists of four main stages: (1) tabular EHR feature construction, (2) graph-based representation learning, (3) multi-task synthetic data generation, and (4) utility-privacy evaluation.

**Figure 1:**
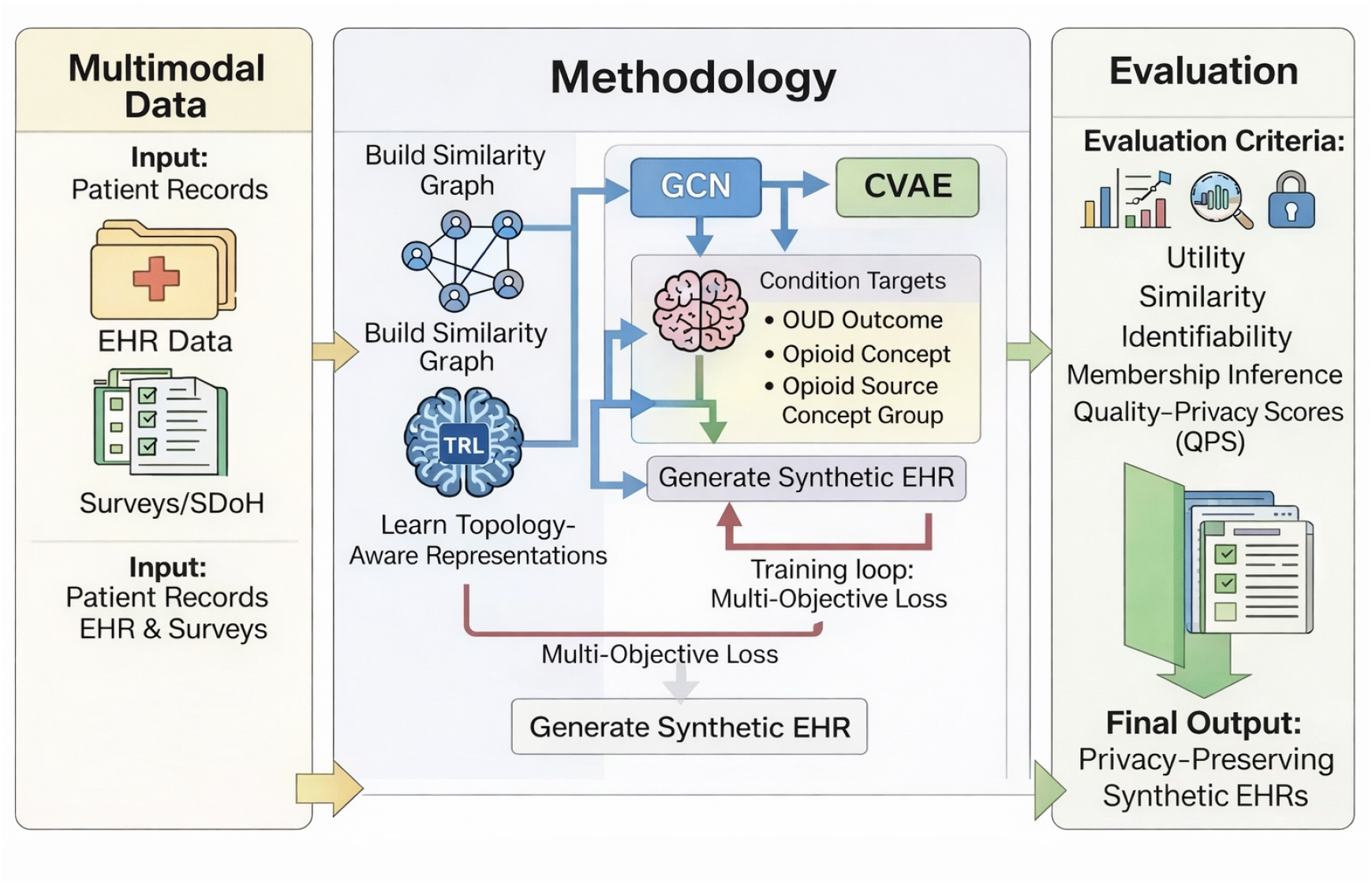
High-level overview of the entire proposed system architecture, illustrating the relationship between core components.

**Figure 2:**
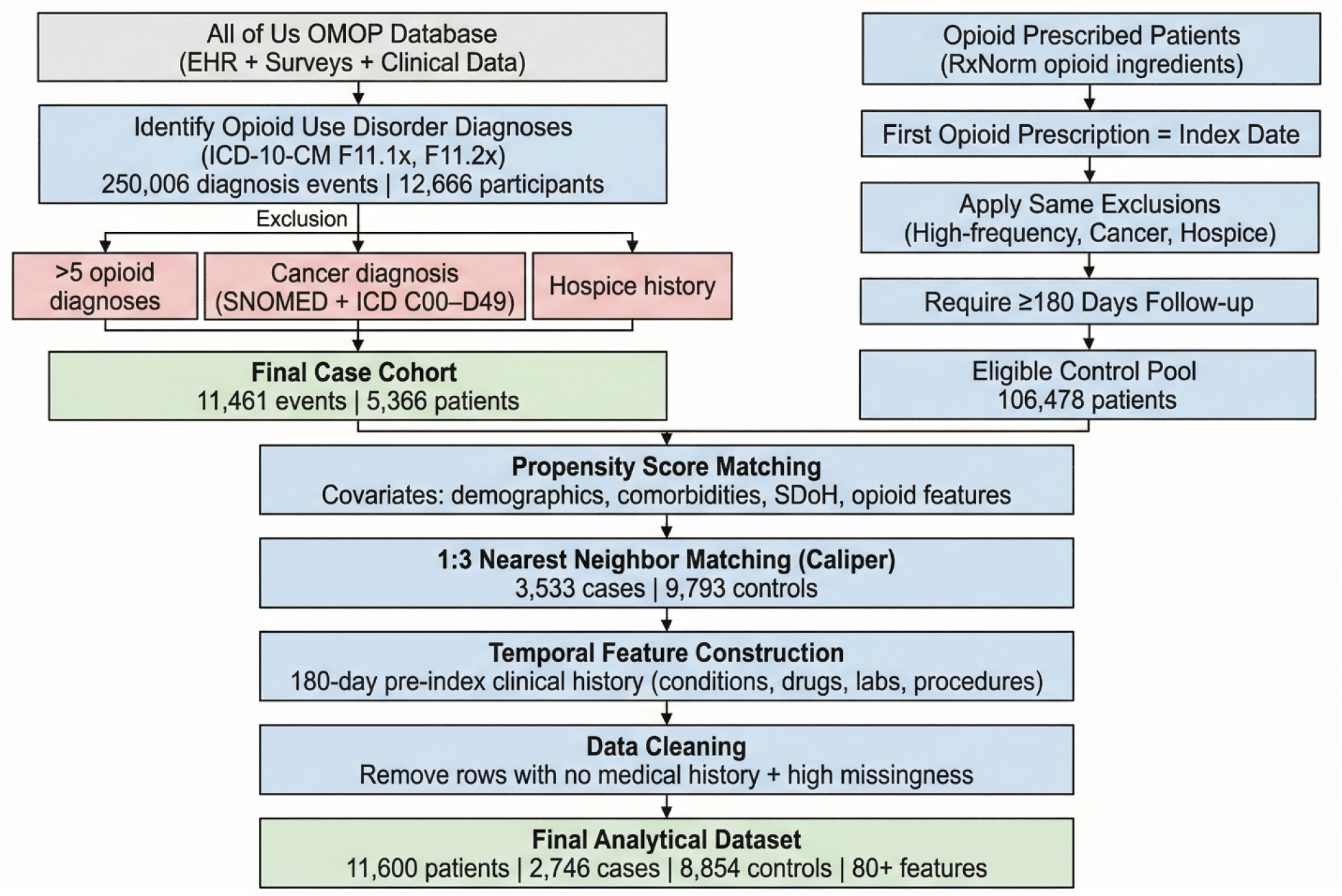
Cohort Selection Flowchart for Opioid Use Disorder Study Using All of Us Data.

**Figure 3:**
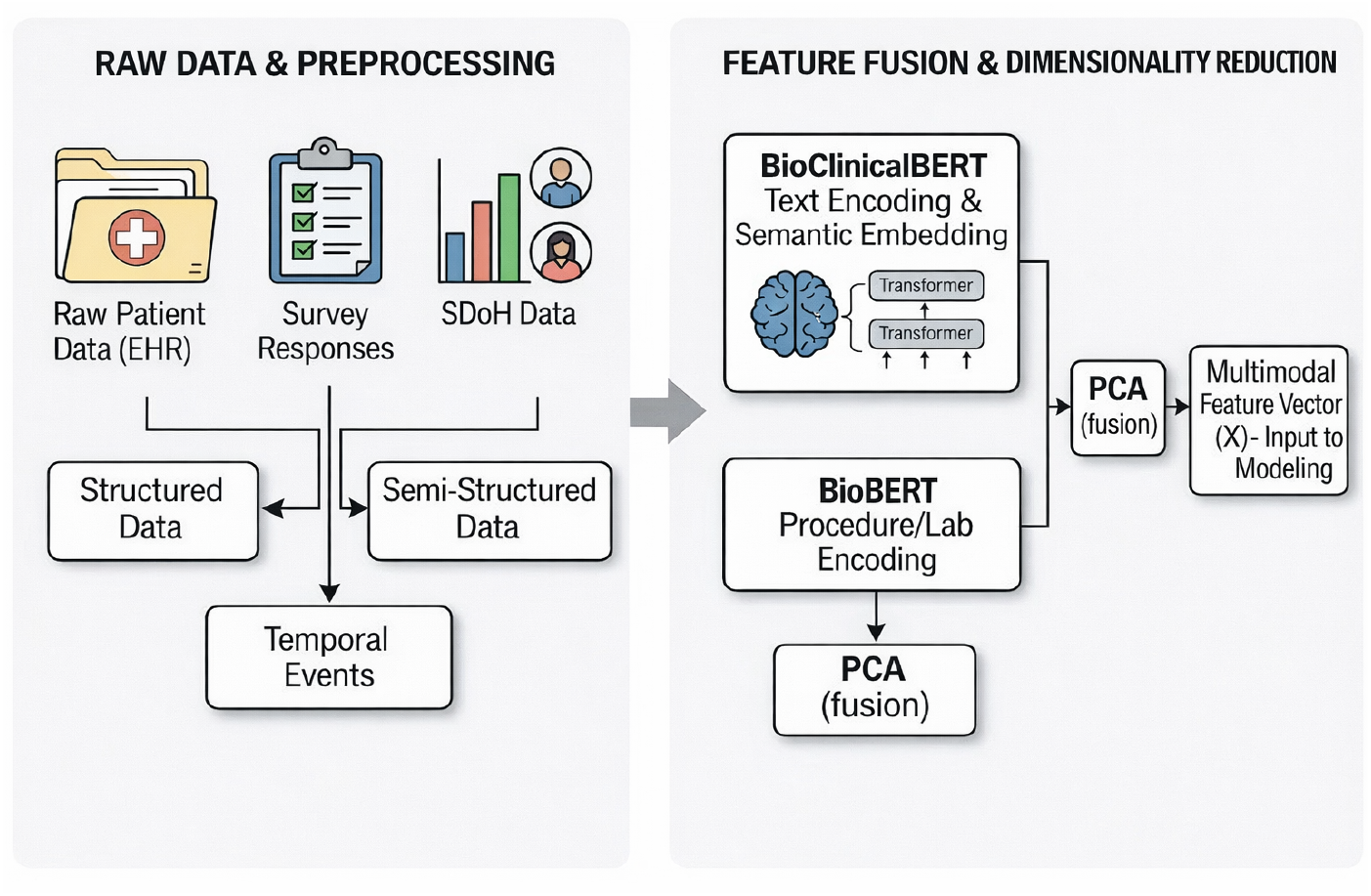
Detailed breakdown of the feature engineering pipeline, including data cleaning, transformation, and selection steps.

**Figure 4:**
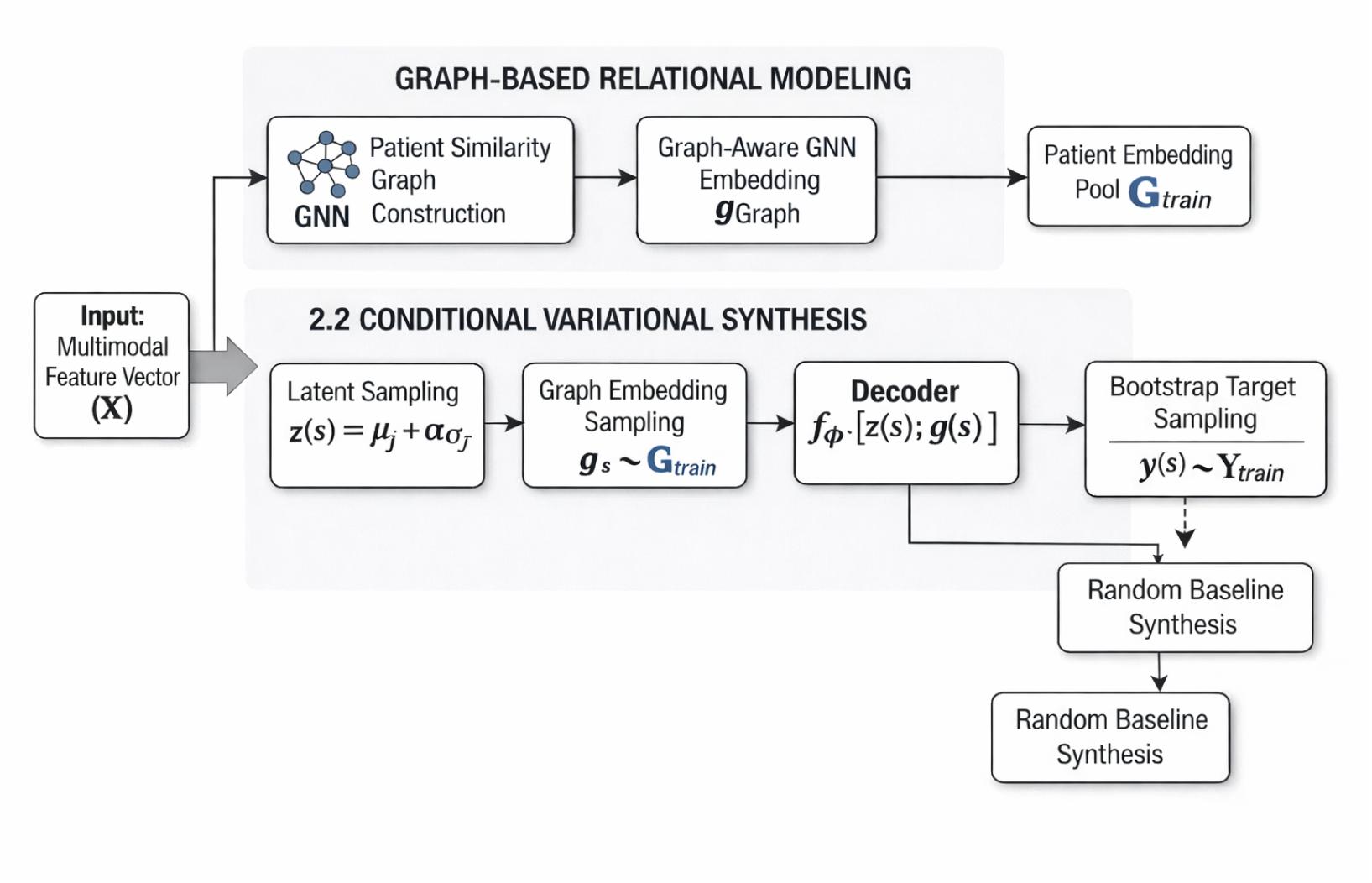
Architecture and information flow within the Multitask Conditional TabGraphSyn

**Figure 5:**
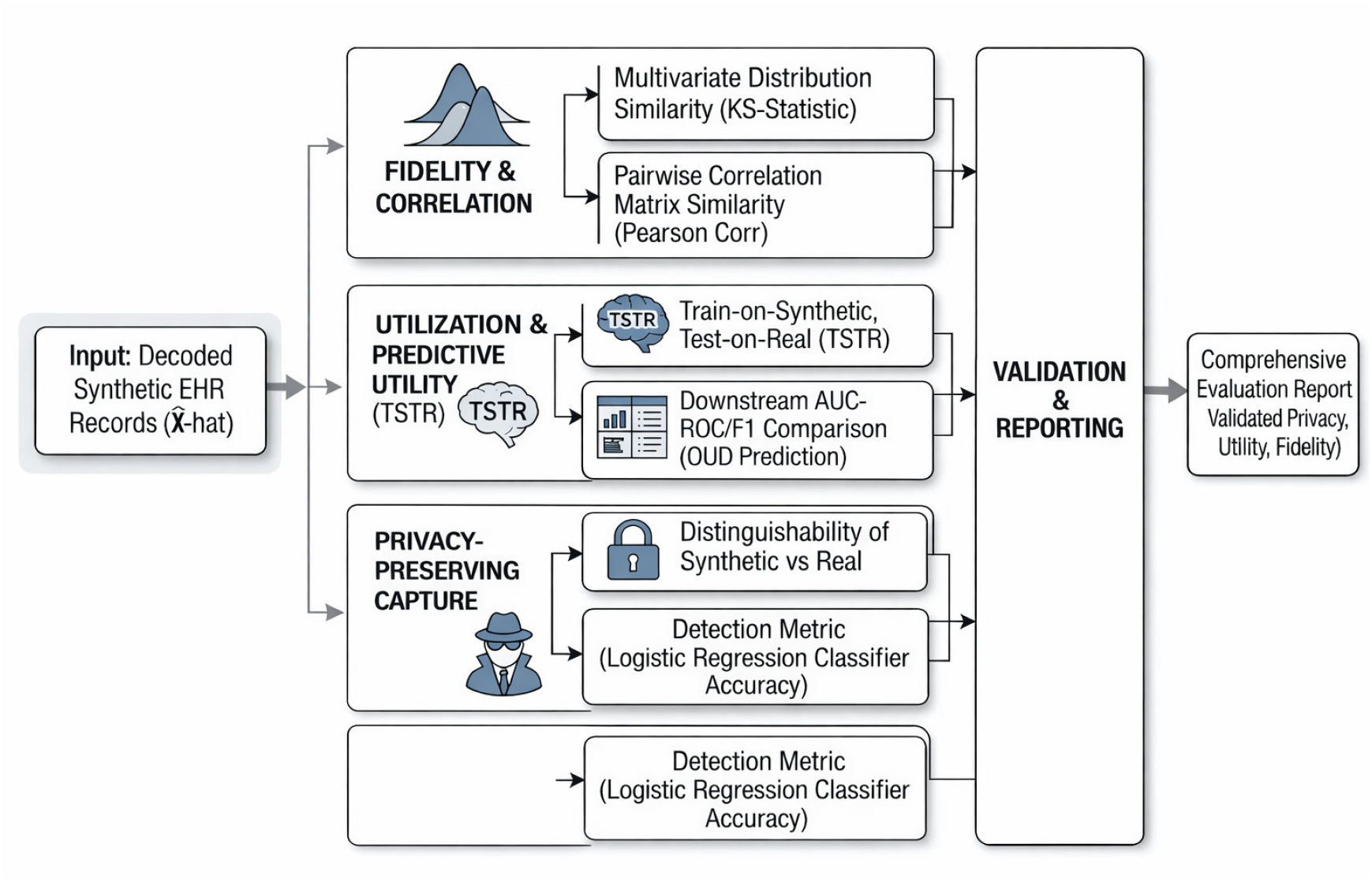
Performance comparison across multiple evaluation metrics (e.g., AUC, F1-Score) contrasted against baseline models.

First, raw EHR data are preprocessed and transformed into a unified tabular representation suitable for synthetic generation and downstream learning. In our setting, the input consists of high-dimensional encoded patient-level features derived from the opioid cohort, along with multiple clinically relevant targets. These targets include *outcome_opioid_abuse_180d, opi-oid_concept_group*, and *opioid_source_concept_group*, which together define the multi-task learning setting.

Second, a patient similarity graph is constructed from the tabular feature space. Each node corresponds to a patient record, and edges connect patients with similar feature profiles. We use this graph structure to capture relational patterns and neighborhood-level dependencies that are not explicitly available in standard tabular modeling. A graph convolutional network (GCN) is then used to learn topology-aware latent embeddings for each patient, providing a compact representation of both feature-level and graph-level structure.

Third, the learned graph embeddings are integrated into a *conditional variational autoencoder* (CVAE) for synthetic data generation. The encoder maps the original tabular features together with graph-derived embeddings into a latent space parameterized by a mean and variance, while the decoder reconstructs synthetic samples conditioned on the graph-aware latent representation. This allows the model to preserve nonlinear feature interactions while remaining guided by the relational structure captured by the graph. To support the multi-task setting, target labels are retained and incorporated during synthetic dataset construction so that the generated data preserve clinically meaningful label-dependent relationships across all three OUD-related tasks.

Finally, synthetic samples are produced by sampling from the learned latent distribution and decoding them back into the original tabular feature space. The generated synthetic records are evaluated against real data and a random baseline under a unified framework that includes distributional similarity, multi-task predictive utility, identifiability risk, membership inference risk, and Quality-Privacy Scores (QPS). We further compare the proposed method against MIIC-SDG to assess whether graph-based deep generative learning provides improved utility-privacy trade-offs in the multi-task opioid use disorder prediction setting.

This integrated framework enables the modeling of complex multivariate dependencies, preserves label-aware structure across multiple prediction tasks, and provides a practical solution for privacy-preserving synthetic EHR generation in clinically sensitive domains.

### 2.2. Data Source and Cohort Construction

This study leverages the NIH *All of Us* Research Program, a large-scale, nationally representative longitudinal cohort that integrates electronic health records (EHR), survey data, and clinical measurements across diverse populations. The dataset follows the Observational Medical Outcomes Partnership (OMOP) common data model, enabling standardized access to clinical domains including diagnoses, procedures, drug exposures, and laboratory measurements. The use of such a harmonized data model facilitates reproducibility and scalable cohort construction across heterogeneous healthcare systems.

#### 2.2.1. Opioid Use Disorder Cohort Definition

The case cohort was defined based on clinically validated opioid use disorder (OUD) diagnoses using ICD-10-CM codes corresponding to opioid abuse and dependence (e.g., F11.1x, F11.2x). All condition occurrences were extracted from the condition_occurrence table, yielding a total of 250,006 diagnosis events across 12,666 unique participants. Each diagnosis event was treated as a longitudinal observation, allowing modeling of repeated clinical encounters and temporal progression.

To improve cohort quality and reduce confounding, several exclusion criteria were applied. Patients with more than five opioid-related diagnoses were excluded to avoid extreme utilization patterns that may bias model learning. Additionally, individuals with any cancer diagnosis were removed using both SNOMED hierarchical concepts (rooted at malignant neoplastic disease) and ICD-10-CM neoplasm codes (C00-D49), as cancer-related opioid use represents a clinically distinct pathway. Patients with any history of hospice care were also excluded, as their care trajectories differ substantially from general populations. After applying these criteria, the final case co-hort consisted of 11,461 diagnosis events across 5,366 unique patients.

#### 2.2.2. Control Cohort Construction via Active Comparator Design

A control cohort was constructed using an active comparator design to ensure clinically meaningful comparison. Specifically, controls were defined as patients who were prescribed opioids but did not develop OUD within a 180-day follow-up window. This design mitigates confounding by indication by ensuring that both cases and controls share similar exposure contexts.

Eligible controls were identified from the drug_exposure table using a curated set of 27 opioid ingredients mapped via RxNorm standard concepts. All descendant drug formulations were included through the OMOP concept_ancestor hierarchy, capturing a comprehensive range of opioid prescriptions. The first opioid prescription was designated as the index date for each control subject.

To ensure temporal validity, only patients with at least 180 days of clinical follow-up after the index date were retained. The same exclusion criteria applied to cases (high-frequency opioid diagnoses, cancer, and hospice history) were also enforced for controls directly within the database query pipeline. This resulted in an initial pool of 106,478 eligible control candidates.

#### 2.2.3. Propensity Score Matching

To reduce selection bias and ensure comparability between cases and controls, propensity score matching (PSM) was performed. A logistic regression model was trained to estimate the probability of OUD onset using a comprehensive set of covariates, including demographic variables (age, sex, race), clinical factors (comorbidities, healthcare utilization), opioid prescription characteristics (ingredient type, route, days supply), and social determinants of health (SDoH).

Matching was conducted using nearest-neighbor matching on the logit-transformed propensity scores with a caliper constraint to limit acceptable distance between matched pairs. A matching ratio of approximately 1:3 (case:control) was achieved, resulting in 3,533 matched cases and 9,793 matched controls. Covariate balance was assessed using standardized mean differences (SMD), with 80% of evaluated covariates achieving acceptable balance (SMD < 0.1), indicating effective reduction of confounding.

#### 2.2.4. Temporal Feature Construction

For each patient, temporal clinical features were derived relative to the index event (first opioid prescription for controls and corresponding reference point for cases). A comprehensive clinical history was constructed by aggregating all events within a 180-day window prior to the index date across four OMOP domains: conditions, drug exposures, measurements, and procedures. These events were encoded as concatenated sequences, capturing rich longitudinal context for each patient.

Additionally, healthcare utilization metrics were computed, including total clinical events and distinct event days. Temporal intervals such as days between last clinical visit and diagnosis, as well as time from prescription to diagnosis, were also derived to capture progression dynamics.

#### 2.2.5. Final Cohort Characteristics

The final dataset consisted of 11,600 patients after data quality filtering, including 2,746 cases and 8,854 matched controls. Each patient record included over 80 features spanning demographics, survey responses, clinical history, prescription data, and derived temporal variables. Rows with completely missing clinical history were removed to ensure sufficient contextual information for downstream modeling.

Despite the high dimensionality, careful preprocessing and cohort design ensured that the dataset preserved clinically meaningful relationships while maintaining robustness for generative modeling. This structured and rigorously curated cohort serves as the foundation for evaluating privacy-preserving synthetic data generation in the context of opioid use disorder prediction.

### 2.3. Feature Engineering and Representation Learning

To effectively model high-dimensional and heterogeneous EHR data, we developed a comprehensive feature engineering pipeline that integrates structured clinical variables with semantically enriched representations derived from longitudinal medical histories. This multimodal representation is designed to capture both explicit clinical attributes and latent contextual information embedded in patient trajectories.

#### 2.3.1. Structured Feature Construction

Structured features were derived from multiple domains within the EHR and survey data. These include demographic attributes (e.g., age, sex, race, and ethnicity), clinical comorbidities (e.g., depression, anxiety, chronic pain), healthcare utilization metrics (e.g., number of encounters and visit frequency), and medication-related variables (e.g., opioid prescription characteristics such as dosage and duration). In addition, survey-derived variables capturing social determinants of health (SDoH), including socioeconomic status, food insecurity, financial strain, and access to healthcare, were incorporated to reflect non-clinical risk factors associated with opioid use disorder.

Categorical variables were transformed using one-hot encoding, while numerical variables were standardized to ensure comparability across features. Missing values were handled using appropriate imputation strategies, ensuring that the resulting feature space remained consistent and suitable for downstream modeling. This structured representation captures patient-level characteristics commonly used in clinical risk modeling, but does not fully reflect the temporal and contextual richness of longitudinal EHR data.

#### 2.3.2. Clinical History Representation via BioClinicalBERT

To incorporate unstructured and longitudinal clinical information, we constructed aggregated textual representations of patient medical histories within a predefined pre-index observation window. Specifically, clinical events were grouped into four major domains: (1) conditions, (2) drug exposures, (3) laboratory measurements, and (4) procedures. For each patient, events within each domain were concatenated into structured text sequences representing their historical clinical trajectory.

These domain-specific sequences were encoded using BioClinicalBERT, a pretrained transformer model fine-tuned on clinical narratives Alsentzer et al. (2019). BioClinicalBERT has been shown to effectively capture biomedical semantics and contextual relationships in clinical text, making it well-suited for representing heterogeneous EHR data. Each text sequence was tokenized and passed through the model, and contextual embeddings were obtained by mean pooling over the final hidden states. This resulted in a dense 768-dimensional representation for each domain.

The use of domain-specific embeddings allows the model to capture nuanced clinical relationships, such as co-occurrence patterns among diagnoses, medication usage trends, and procedural sequences, which are not easily represented using traditional tabular encoding methods. Moreover, these embeddings provide a semantically meaningful compression of highdimensional clinical information into a fixed-length vector representation.

#### 2.3.3. Multimodal Feature Fusion

The final patient representation was constructed by concatenating structured features with the four domain-specific BioClinicalBERT embeddings, resulting in a high-dimensional multimodal feature vector. This unified representation combines interpretable clinical variables with latent semantic features, enabling the model to leverage both explicit and implicit information in the data.

To address the curse of dimensionality and improve computational efficiency, dimensionality reduction was subsequently applied using principal component analysis (PCA), projecting the combined feature space into a lower-dimensional latent representation while preserving the majority of variance in the data. This compressed representation serves as the input to subsequent graph construction and generative modeling stages.

By integrating structured clinical variables with contextual embeddings derived from longitudinal EHR data, the proposed representation learning framework captures complex multivariate dependencies and enhances the model’s ability to generate realistic and clinically meaningful synthetic patient records.

### 2.4. Graph Construction and Relational Modeling

To capture relational dependencies that extend beyond independent tabular representations, we construct a patient-level similarity graph that explicitly models interactions among individuals in the cohort. This graph-based formulation enables the learning of structured dependencies that are not directly observable through conventional feature engineering approaches.

#### 2.4.1. Patient Similarity Graph Construction

Given the multimodal feature representation described in the previous section, we construct a *k*-nearest neighbor (k-NN) graph over patients in the latent feature space. Each node in the graph corresponds to an individual patient, while edges represent similarity relationships computed using distance metrics in the learned feature space. Specifically, we compute pairwise distances between patient embeddings and connect each node to its *k* nearest neighbors, resulting in a sparse adjacency structure that captures local neighborhood relationships.

The resulting graph can be represented as *G = (V, E)*, where *V* denotes the set of patients and *E* denotes the set of similarity-based edges. The adjacency matrix is subsequently symmetrized and normalized to ensure stable downstream learning. This graph construction allows the model to encode population-level structure, where patients with similar clinical profiles are grouped together, facilitating the propagation of information across related individuals.

#### 2.4.2. Graph-Aware Representation Learning

Building upon the constructed patient graph, we learn graph-aware embeddings that integrate both node-level attributes and neighborhood context. This step enables the model to capture higher-order interactions and shared patterns across patients, which are particularly important in clinical settings where disease trajectories and treatment responses exhibit cohort-level similarities.

Unlike traditional tabular approaches that treat samples as independent observations, the graph-based formulation allows information to be aggregated across similar patients, effectively enhancing representation quality in sparse or noisy regions of the data space. This is especially beneficial for healthcare datasets, where individual patient records may be incomplete or heterogeneous.

#### 2.4.3. Relation to Prior Graph-Based Approaches

Graph-based modeling of healthcare data has been explored through both probabilistic dependency learning and graph neural representation learning. A notable recent example is MIIC-SDG, which uses multivariate information theory and Bayesian network modeling to reconstruct feature-level dependency structures and generate synthetic samples under a probabilistic graphical framework Sella et al. (2025). This line of work is valuable because it explicitly models conditional dependence patterns and introduces a principled quality-privacy evaluation framework based on Quality-Privacy Scores (QPS). However, MIIC-SDG primarily operates at the level of feature dependencies and is less flexible in capturing complex nonlinear interactions across high-dimensional patient records. In addition, it is not naturally designed to preserve relationships across multiple downstream prediction targets simultaneously.

Patient-level graph-based generative methods such as TabGraphSyn provide a complementary perspective. Rather than learning only feature-to-feature dependencies, these approaches construct similarity graphs among patients and use graph neural networks to learn topology-aware embeddings that encode both individual attributes and neighborhood context. This formulation is particularly well suited for EHR data, where clinically meaningful structure often emerges from similarities among patients with related histories, comorbidities, and treatment trajectories. In its original form, however, TabGraphSyn is primarily a single-task synthetic generation framework and does not explicitly target preservation of multiple clinically relevant labels.

Our proposed **Multi-Task TabGraphSyn** builds on this patient-graph paradigm while extending it to a multi-task opioid use disorder setting. Specifically, we retain the graph-based relational foundation of TabGraphSyn, use a graph convolutional network (GCN) to learn patient embeddings, and integrate these embeddings into a conditional variational autoencoder (CVAE) for synthetic data generation. Compared with MIIC-SDG, the proposed framework emphasizes patient-level relational learning and nonlinear representation capacity; compared with prior TabGraphSyn formulations, it explicitly preserves joint label-dependent structure across multiple OUD-related targets. This makes the method better aligned with our experimental objective of generating synthetic EHR data that are not only statistically realistic, but also useful for multi-task downstream prediction under a rigorous utility-privacy evaluation framework.

#### 2.4.4. Role of Graph Structure in Generative Modeling

In the proposed framework, the patient similarity graph acts as a structural prior that informs the synthetic data generation process. By connecting patients with similar encoded feature profiles, the graph captures cohort-level organization that is not directly represented by independent tabular observations. This is especially important in clinical data, where relevant patterns often arise from shared comorbidities, medication histories, utilization behavior, and other population-level similarities rather than from isolated features alone.

The graph convolutional network uses this adjacency structure to propagate information across neighboring patients and produce topology-aware embeddings. These embeddings summarize both local patient attributes and neighborhood context, thereby serving as a relationally informed latent representation of the cohort. The downstream CVAE then conditions on these graph-derived embeddings during encoding and decoding, so that synthetic samples are generated from a latent space that already reflects patient similarity structure.

This design offers two important advantages. First, it improves fidelity by encouraging generated samples to remain consistent with observed cohort-level patterns in the real data. Second, it helps preserve clinically meaningful label-dependent relationships in the multi-task setting, since patients with similar feature and graph profiles are more likely to share related opioid outcomes and concept group patterns. In this sense, the graph does not replace the generative model; rather, it regularizes and enriches it by embedding each patient record in a broader relational context.

Overall, the integration of graph construction and graph-aware representation learning provides a principled bridge between high-dimensional tabular EHR features and multi-task synthetic generation. It enables Multi-Task TabGraphSyn to model dependencies that are difficult to capture through purely feature-based probabilistic models such as MIIC-SDG or through tabular generative models that ignore inter-patient structure.

### 2.5. Conditional Generative Model

To model the high-dimensional distribution of EHR data while preserving clinically meaningful relational structure, we employ a graph-conditioned variational generative model. Unlike our earlier diffusion-based formulation, the proposed **Multi-Task TabGraphSyn** adopts a simpler and more stable architecture that combines a graph convolutional encoder with a conditional variational autoencoder (CVAE). This design is better aligned with the tabular nature of the data and supports efficient synthetic generation under a multi-task evaluation framework.

#### 2.5.1. Graph-Aware Latent Representation

Let **x**_***i***_ ∈ ℝ^*d*^ denote the encoded tabular feature vector for patient *i*, where the feature space includes one-hot encoded categorical variables, numerical clinical attributes, and other derived patient-level representations after imputation and standardization. From these patient representations, we construct a *k*-nearest neighbor graph over the training set. The resulting adjacency matrix is symmetrized and degree-normalized to obtain a graph operator **Ã**.

A graph convolutional encoder is then used to learn topology-aware patient embeddings. Given the input feature matrix **X** and normalized adjacency matrix **Ã**, the encoder computes

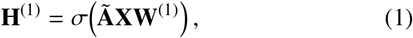

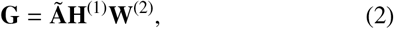

where σ(·) denotes a nonlinear activation function, **W**^(1)^ and **W**^(2)^ are learnable parameters, and 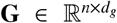 contains the graph-aware embeddings for all patients. These embeddings summarize both the original patient attributes and local neighborhood structure, thereby encoding cohort-level relational information that is absent in standard independent tabular models.

#### 2.5.2. Conditional Variational Autoencoder

The graph-derived embedding ***g***_***i***_ for patient *i* is concatenated with the standardized tabular feature vector ***x***_***i***_ and passed to a variational encoder:

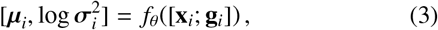

where *f*_*θ*_(·) is implemented as a multilayer perceptron and [·; ·] denotes concatenation. The encoder parameterizes a latent Gaussian distribution,

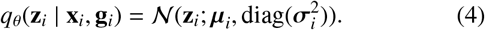

A latent sample is then obtained via the reparameterization trick:

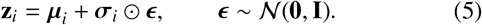

The decoder reconstructs the original feature vector conditioned on both the latent variable and the graph embedding:

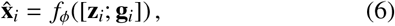

where *f*_*ϕ*_ (·) is a neural decoder. This conditional design allows the generator to produce samples that are not solely driven by a low-dimensional latent code, but are also informed by graph-aware patient context.

#### 2.5.3. Multi-Task Conditioning Strategy

The proposed method is designed for a multi-task opioid use disorder setting with three targets:

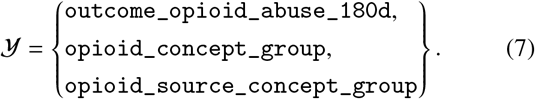

Rather than directly injecting task labels into the decoder as explicit conditioning vectors, we preserve multi-task structure during synthetic dataset construction. After generating synthetic feature vectors, target variables are sampled from the empirical joint training distribution through bootstrap resampling of the multi-task label matrix. This strategy ensures that the synthetic dataset retains realistic task marginals and label cooccurrence patterns observed in the real data, while allowing the generative model itself to focus on learning faithful feature distributions conditioned on graph-aware structure.

This design is consistent with our experimental objective: evaluating whether synthetic data generated by Multi-Task TabGraphSyn can support downstream multi-output classification on held-out real data. Thus, multi-task preservation is assessed operationally through train-on-synthetic, test-on-real evaluation using the three opioid-related targets.

#### 2.5.4. Synthetic Sample Generation

After training, synthetic records are generated by first encoding training samples to obtain latent parameters 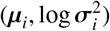 and then sampling latent vectors from these learned posterior regions. Specifically, synthetic latent codes are generated as

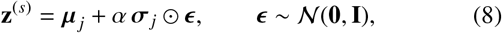

where *j* indexes a sampled training instance and *α* controls the sampling noise scale. A graph embedding **g**^(*s*)^ is sampled from the pool of graph-aware embeddings, and the synthetic feature vector is decoded as

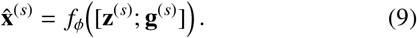

The decoded features are then transformed back to the original encoded feature space using the inverse of the standardization pipeline. Binary variables are thresholded to valid values, and continuous variables are clipped to observed training ranges to improve plausibility. Finally, synthetic task labels are appended through bootstrap sampling from the training multitask target distribution.

#### 2.5.5. Model Advantages

The proposed conditional generative model provides several advantages for privacy-preserving EHR synthesis. First, the graph convolutional encoder captures inter-patient similarity structure that is ignored by purely tabular generative models. Second, the CVAE offers a stable and computationally efficient mechanism for learning nonlinear data distributions in high-dimensional encoded feature spaces. Third, the framework naturally supports downstream multi-task evaluation by generating synthetic cohorts that preserve the statistical structure required for simultaneous prediction of opioid abuse outcome, opioid concept group, and opioid source concept group. Relative to MIIC-SDG, which emphasizes feature-level dependency reconstruction, Multi-Task TabGraphSyn emphasizes patient-level relational learning and nonlinear generative capacity, thereby offering a complementary and often stronger utility-privacy trade-off in our experimental setting.

### 2.6. Training Objective and Optimization

The proposed Multi-Task TabGraphSyn framework is trained in two stages: graph representation learning and conditional variational generation. This staged optimization improves training stability and separates the learning of relational structure from the learning of the generative feature distribution.

#### 2.6.1. Graph Encoder Objective

The graph convolutional encoder is trained on the patient similarity graph using a smoothness-oriented unsupervised objective. Let **G** denote the matrix of graph embeddings produced by the GCN. We encourage neighboring patients to have similar embeddings by minimizing a graph smoothness loss:

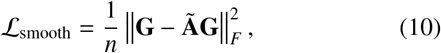

where **Ã** is the normalized adjacency matrix and || ·||_***F***_ denotes the Frobenius norm. To prevent embedding magnitudes from growing without bound, we add a small regularization term:

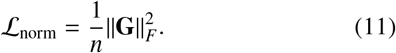

The final GCN training objective is

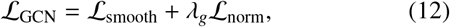

where *λ*_***g***_ is a small regularization coefficient. This objective promotes topology-aware latent representations that respect local graph structure while remaining numerically stable.

#### 2.6.2. Variational Reconstruction Objective

After learning graph embeddings, the conditional VAE is trained using paired inputs (**x**_***i***_, **g**_***i***_). The loss combines a reconstruction term and a Kullback-Leibler divergence term. The reconstruction loss is defined as mean squared error between the standardized input and reconstruction:

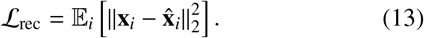

The KL regularization encourages the posterior to remain close to a standard Gaussian prior:

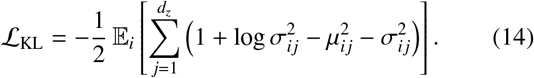

The total CVAE objective is

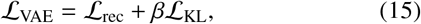

where *β* controls the strength of regularization. This objective encourages accurate feature reconstruction while maintaining a smooth latent space from which synthetic samples can be generated.

#### 2.6.3. Optimization Procedure

The GCN and CVAE are optimized separately using Adam. For the GCN, training proceeds over the graph-structured training set until the graph smoothness objective converges, with early stopping based on the best observed training loss. Once the graph encoder is fixed, graph embeddings for validation and test samples are obtained through an out-of-sample projection learned from the training embeddings. These projected embeddings allow the CVAE to condition consistently on graph-aware representations across dataset splits.

The CVAE is then trained using mini-batches of feature vectors and graph embeddings. Early stopping is applied using validation loss to reduce overfitting. In practice, this two-stage training scheme was found to be more stable than the previously explored diffusion-based alternative and better suited for the encoded opioid cohort used in this study.

#### 2.6.4. Connection to Multi-Task Evaluation

Although the generative objective itself is unsupervised with respect to direct classification loss, the model is explicitly developed for a multi-task synthetic data setting. After training, the quality of the learned synthetic distribution is assessed by training a `MultiOutputClassifier` on synthetic data and evaluating it on held-out real test data across all three target tasks. Performance is summarized using macro-averaged accuracy, precision, recall, F1-score, ROC-AUC, and AUPRC across tasks. In addition, the model is evaluated using identifiability risk, membership inference risk, normalized quality scores, and composite QPS and MetaQPS measures aligned with the MIIC-SDG evaluation framework.

#### 2.6.5. Practical Rationale

This optimization strategy reflects a deliberate methodological choice. The earlier graph-latent diffusion formulation was more complex and introduced additional instability without yielding stronger results in our setting. In contrast, the final Multi-Task TabGraphSyn design offers a more parsimonious architecture with clear graph conditioning, efficient training, and direct compatibility with the MIIC-style privacy-utility evaluation pipeline. This makes it more appropriate for the current study, whose central contribution is not a new diffusion mechanism, but a robust multi-task extension of TabGraphSyn for opioid use disorder prediction and privacy-preserving synthetic EHR generation.

### 2.7. Synthetic Data Generation

Once the proposed Multi-Task TabGraphSyn model is trained, synthetic EHR data are generated through the graph-conditioned variational latent space rather than through a reverse diffusion process. The generation procedure is designed to preserve feature-level realism, cohort-level relational structure, and multi-task label distributions relevant to opioid use disorder.

Let 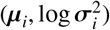denote the latent parameters learned by the conditional variational encoder for training instance *i*, conditioned on both the tabular feature vector **x**_***i***_ and its graph-aware embedding **g**_***i***_. To generate a synthetic sample, we first select a training instance index *j* by bootstrap sampling and draw a latent vector from its local posterior neighborhood:

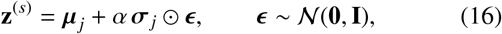

where *α* is a noise-scaling parameter controlling the variability of the generated samples.

In parallel, a graph-aware embedding is sampled from the pool of learned patient embeddings:

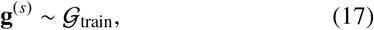

where 𝒢_train_ denotes the empirical distribution of graph-derived embeddings obtained from the training and validation data. The synthetic feature vector is then decoded conditionally as

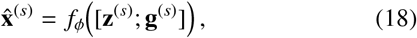

where *f*_*ϕ*_ (·) is the trained decoder and [·; ·] denotes vector concatenation.

The decoder output is produced in the standardized encoded feature space. To obtain synthetic data in the original modeling domain, the decoded vectors are transformed back using the inverse of the preprocessing pipeline:

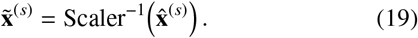

Post-processing is then applied to enforce valid feature types. Binary or binary-like variables are thresholded to {0,1}, while continuous features are clipped to the observed minimum-maximum ranges of the real training data in order to reduce implausible extrapolation:

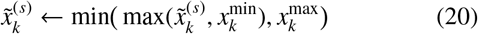

for each continuous feature *k*.

To support the multi-task setting, synthetic target variables are appended after feature generation. Rather than predicting labels directly from the decoder, we sample the multi-task target tuples from the empirical training distribution using bootstrap resampling:

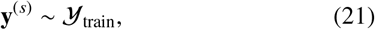

where **y**^(*s*)^ contains the three opioid-related targets: *out-come_opioid_abuse_180d, opioid_concept_group*, and *opi-oid_source_concept_group*. This strategy preserves realistic label frequencies and co-occurrence patterns observed in the original cohort while keeping the feature generator focused on modeling the patient feature distribution.

The synthetic cohort size can be configured to match either the full real training-validation cohort or only the training subset. In our experiments, the synthetic sample size is set to match the real cohort size used for comparison, enabling fair downstream evaluation and privacy analysis.

In addition to the proposed synthetic generator, we construct a random baseline synthetic dataset for normalization in the MIIC-style evaluation framework. For this baseline, binary features are sampled from Bernoulli distributions estimated from empirical feature means, whereas continuous features are sampled uniformly within observed training ranges. The same bootstrap strategy is used to assign multi-task labels. This baseline provides a reference point for normalized quality and privacy scores such as *Q*_mi_, *Q*_corr_, *Q*_wasserstein_, and the downstream QPS and MetaQPS metrics.

Overall, this generation process produces fully synthetic patient records that reflect the learned graph-aware latent structure of the cohort, preserve multi-task target distributions relevant to opioid use disorder, and support rigorous evaluation of utility, fidelity, and privacy. The resulting synthetic data can be used for downstream benchmarking, train-on-synthetic test-on-real experiments, and privacy-aware data sharing in sensitive clinical settings.

## 3. Experiments and Evaluation

### 3.1. Experimental Setup

We evaluated the proposed Multi-Task TabGraphSyn framework on the opioid use disorder cohort derived from the NIH *All of Us* Research Program. The analysis focused on three clinically relevant prediction targets: (i) opioid abuse within 180 days, treated as the primary binary outcome, (ii) opioid concept group, and (iii) opioid source concept group, both formulated as multi-class targets. This multi-task formulation was chosen to assess whether synthetic data could preserve not only overall feature distributions, but also the joint label structure required for clinically meaningful downstream prediction.

After cohort construction and feature preprocessing, the data were partitioned into training, validation, and test subsets. The initial split reserved 20% of the cohort as a held-out test set, and an additional 10% of the remaining development set was used for validation. Stratification was performed with respect to the primary opioid abuse outcome to preserve class balance across splits. All synthetic data generators were trained exclusively on the development data, and all final utility evaluations were conducted on the held-out real test set to avoid information leakage.

A consistent preprocessing pipeline was applied throughout the study. Categorical variables were converted to one-hot representations, missing values were imputed using median imputation, and all features were standardized prior to graph construction and model training. The same encoded feature space was used for both model development and evaluation to ensure comparability between real and synthetic cohorts. For the proposed method, graph construction, graph embedding learning, variational generation, and synthetic sampling were all performed within each experimental split.

To assess robustness, all experiments were repeated over four random seeds (42, 43, 44, and 45). For each seed, we trained the model, generated a full synthetic cohort, and computed all evaluation metrics independently. Final reported results were then summarized across seeds using the mean and standard deviation.

The evaluation protocol was designed to match the MIIC-SDG framework as closely as possible. We assessed synthetic data quality from multiple perspectives: downstream predictive utility, feature-level and multivariate distributional similarity, identifiability risk, membership inference risk, normalized quality and privacy scores, and composite Quality-Privacy Scores (QPS). In addition, a random synthetic baseline was generated in each run to normalize quality and privacy measures in the same manner used by the MIIC-style evaluation pipeline.

For downstream utility, classifiers were trained under three settings: real-train/real-test, synthetic-train/real-test, and random-train/real-test. Multi-task prediction was implemented using a `MultiOutputClassifier` with logistic regression base learners, and performance was summarized using macro-averaged accuracy, precision, recall, Fl-score, ROC-AUC, and area under the precision-recall curve across the three tasks. This evaluation strategy allowed us to quantify how well the synthetic data preserved the information necessary for simultaneous prediction of opioid abuse risk and opioid-related concept groupings on unseen real patients.

### 3.2. Baseline Method

We compared the proposed method against a single baseline, MIIC-SDG, which served as the primary reference model throughout the study.

- **MIIC-SDG**: MIIC-SDG is a recent synthetic data generation framework based on multivariate information theory and Bayesian network modeling Sella et al. (2025). The method reconstructs dependency structures among variables using conditional mutual information and then generates synthetic data through probabilistic sampling from the learned graphical model. In addition to providing a strong mechanism for preserving multivariate statistical associations, MIIC-SDG introduced the Quality-Privacy Score (QPS) framework for jointly assessing synthetic data fidelity and privacy risk.

MIIC-SDG was selected as the sole baseline because it is the most relevant comparator for the present work. First, it addresses the same core problem of privacy-preserving synthetic data generation for clinical datasets. Second, it provides a fundamentally different modeling strategy, emphasizing feature-level probabilistic dependency learning rather than patient-level graph representation learning. This contrast allows a clearer examination of whether graph-aware deep generative modeling offers advantages over information-theoretic graphical synthesis in the multi-task opioid use disorder setting. Third, the evaluation framework used in this study was intentionally aligned with the MIIC-SDG protocol, including normalized quality metrics, privacy metrics, QPS, and MetaQPS.

As a result, MIIC-SDG provides not only a methodological baseline but also an evaluation anchor for interpreting the observed utility-privacy trade-offs.

Accordingly, all results reported in this work compare MultiTask TabGraphSyn directly against MIIC-SDG under a unified experimental design. This enables a focused and fair assessment of whether the proposed graph-based multi-task generative framework improves the balance between predictive utility, distributional fidelity, and privacy preservation in synthetic EHR generation for opioid use disorder research.

### 3.3. Evaluation Metrics

Evaluating synthetic clinical data requires a framework that jointly measures fidelity to the original data distribution, usefulness for downstream prediction, and vulnerability to privacy attacks. To enable a direct comparison with MIIC-SDG, we adopted the same broad evaluation philosophy and implemented a matched protocol around five complementary dimensions: reconstruction and feature-level similarity, multivariate distributional quality, multi-task predictive utility, privacy risk, and normalized quality-privacy trade-off scores.

#### 3.3.1. Reconstruction and Feature-Level Similarity

We first quantified how closely the generated synthetic records resembled real patient records in the standardized feature space. Following generation, we computed proxy matching statistics between sampled real and synthetic observations, including mean squared error (MSE), mean absolute error (MAE), and average cosine similarity. Let **x**^(*r*)^ and **x**^(*s*)^ denote matched real and synthetic feature vectors, respectively. The reconstruction-oriented discrepancies are defined as

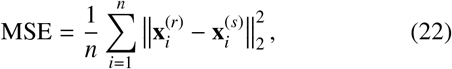

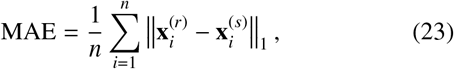

and cosine similarity was computed pairwise and averaged over sampled record pairs. Lower MSE and MAE, together with higher cosine similarity, indicate closer agreement between real and synthetic observations.

To further assess first-order feature fidelity, we measured the average absolute gap in feature means and feature standard deviations between the real training cohort and the synthetic cohort:

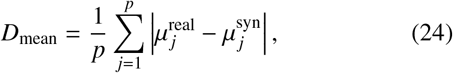

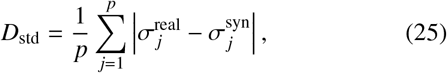

where *p* is the number of evaluated features. These metrics provide an interpretable summary of how well the synthetic data preserve marginal feature behavior.

#### 3.3.2. Multivariate Distributional Quality

Because synthetic EHR quality depends on preserving relationships across variables rather than only univariate marginals, we evaluated multivariate structure using three complementary distances.

First, we measured **correlation distance** by computing correlation matrices for selected real and synthetic feature subsets and taking the mean absolute element-wise difference:

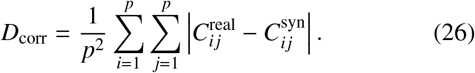

Second, we evaluated **mutual information distance** to capture dependence patterns that extend beyond linear association. Continuous variables were discretized by quantile binning, binary-like variables were preserved as categorical states, and pairwise mutual information matrices were computed for real and synthetic data. The resulting distance was

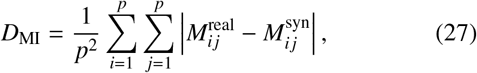

where *M*_*ij*_ denotes the mutual information between variables *i* and *j*.

Third, we computed a **global Wasserstein distance** after projecting real and synthetic samples into a low-dimensional principal component space. Let 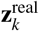 and 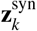’ denote the projected scores along principal component *k*. The Wasserstein distance was averaged across retained components:

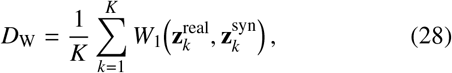

where *W*_1_ (·, ·) is the first-order Wasserstein distance and *K* is the number of principal components used in evaluation. Lower values of *D*_corr_, *D*_MI_, and *D*_W_ indicate better preservation of multivariate structure.

#### 3.3.3. Multi-Task Predictive Utility

To assess whether the synthetic data preserve clinically meaningful signal for downstream learning, we adopted a train-on-synthetic, test-on-real evaluation scheme in a multi-task setting. Specifically, we trained a `MultiOutputClassifier` with logistic regression base learners on synthetic data and evaluated performance on the held-out real test set. The same classifier was also trained on real data and on the random synthetic baseline for comparison.

The three prediction tasks were: opioid abuse within 180 days, opioid concept group, and opioid source concept group. For each task, we computed accuracy, macro-averaged precision, macro-averaged recall, macro-averaged F1-score, macro-averaged one-vs-rest ROC-AUC, and macro-averaged area under the precision-recall curve (AUPRC). Task-specific metrics were then averaged to obtain overall multi-task utility summaries. Let 𝒯 denote the set of tasks and let *m*_*t*_ be a metric computed on task *t*. The macro-average across tasks was defined as

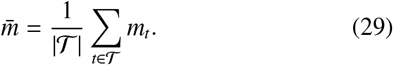

We report both the synthetic-train/real-test utility and the corresponding real-train/real-test reference performance. To quantify the loss in downstream usefulness, we additionally computed utility gaps for macro F1 and macro ROC-AUC:

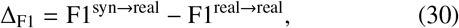

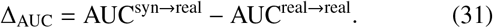

#### 3.3.4. Privacy Risk Assessment

We evaluated privacy from two attack-oriented perspectives: identifiability and membership inference.

##### Identifiability

Identifiability measures how often a real patient has a synthetic neighbor that is at least as close as the patient’s nearest real neighbor. To make this criterion sensitive to rare or low-entropy variables, features were first weighted by inverse entropy. Let *d*_***rr***_(*i*) denote the distance from real sample *i* to its nearest other real sample, and let *d*_***rs***_(*i*) denote the distance from the same real sample to its nearest synthetic sample. A real sample was considered identifiable if

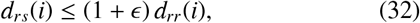

where *ϵ* was set to zero in our experiments. The identifiability score was then defined as

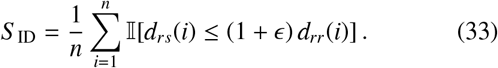

Lower identifiability implies better privacy, and we also report the corresponding privacy quantity 1 - *S*_ID_.

##### Membership inference

Membership inference evaluates whether an attacker can determine if a real sample belonged to the training set based on its distance to synthetic samples. We compared member samples (training records) against non-member samples (held-out test records). After projecting real and synthetic data into a shared principal component space, nearest-neighbor distances from members and non-members to the synthetic set were computed. These distances were converted into attack scores and summarized using ROC-AUC:

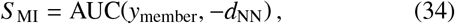

where *y*_member_indicates membership status and *d*_NN_ is the nearest-synthetic distance. A value near 0.5 indicates weak attack success, while larger values indicate stronger privacy leakage. We also report the corresponding privacy quantity 1 - *S* _MI_ and the balanced accuracy of the thresholded attack.

#### 3.3.5. Normalized Quality and Privacy Scores

To make quality and privacy scores directly comparable across methods, we normalized both using a random synthetic baseline generated from empirical marginal distributions and training-range sampling. For any quality distance *D*_syn_ and the corresponding random-baseline distance *D*_rand_, the normalized quality score was defined as

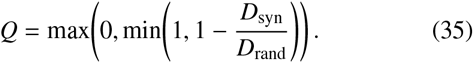

We computed three such quality measures:

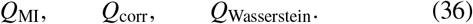

Similarly, for privacy attack scores *S* _syn_ and *S* _rand_, normalized privacy was defined as

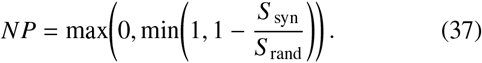

This yielded two normalized privacy measures:

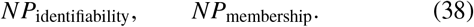

Under this normalization, values closer to one indicate stronger improvement over the random baseline, while values near zero indicate little advantage.

#### 3.3.6. Quality-Privacy Scores (QPS) and MetaQPS

Following the MIIC-SDG evaluation framework, we combined normalized quality and privacy scores using harmonic means to capture the trade-off between utility-preserving realism and privacy protection. For a given normalized quality score *Q* and normalized privacy score *NP*, the corresponding Quality-Privacy Score was

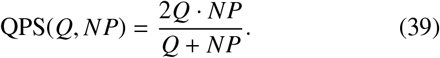

We report six pairwise QPS measures obtained by combining each of the three normalized quality scores with the two normalized privacy scores:

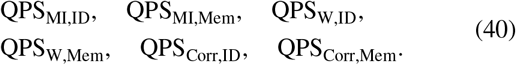

To summarize the overall balance between quality and privacy, we further computed two aggregate scores. First, an arithmetic-mean variant was formed by averaging the normalized quality scores and privacy scores separately and then taking their harmonic mean:

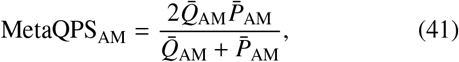

where

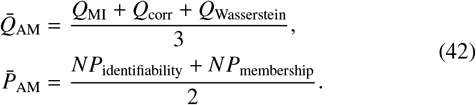

Second, a harmonic-mean variant was computed by first aggregating quality and privacy internally through harmonic means and then combining them:

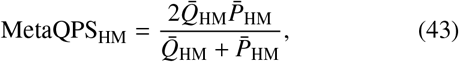

where

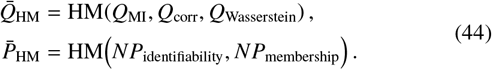

These composite measures provide a concise summary of overall method performance under the joint objective of generating synthetic data that are both useful and privacy-preserving.

### 3.3.7. Summary of the Evaluation Framework

Taken together, the evaluation framework assesses the proposed method from three complementary perspectives. First, it measures fidelity through reconstruction-oriented statistics, feature moment alignment, and multivariate structural similarity. Second, it measures utility through multi-task train-on-synthetic, test-on-real prediction across opioid-related outcomes. Third, it measures privacy through identifiability and membership inference attacks, and integrates these with normalized quality scores through QPS and MetaQPS. This matched protocol enables a direct and interpretable comparison between Multi-Task TabGraphSyn and MIIC-SDG under a unified utility-privacy framework.

## 4. Results

This study compared Multi-Task TabGraphSyn against MIIC-SDG under a unified evaluation framework designed to assess synthetic EHR data from the perspectives of downstream utility, distributional fidelity, privacy risk, and overall quality-privacy trade-off. Several important findings emerge from the results. As summarized in Table 1, Multi-Task TabGraphSyn achieved a stronger overall balance between utility, fidelity, privacy, and composite quality-privacy trade-off metrics than MIIC-SDG.

**Table 1:**
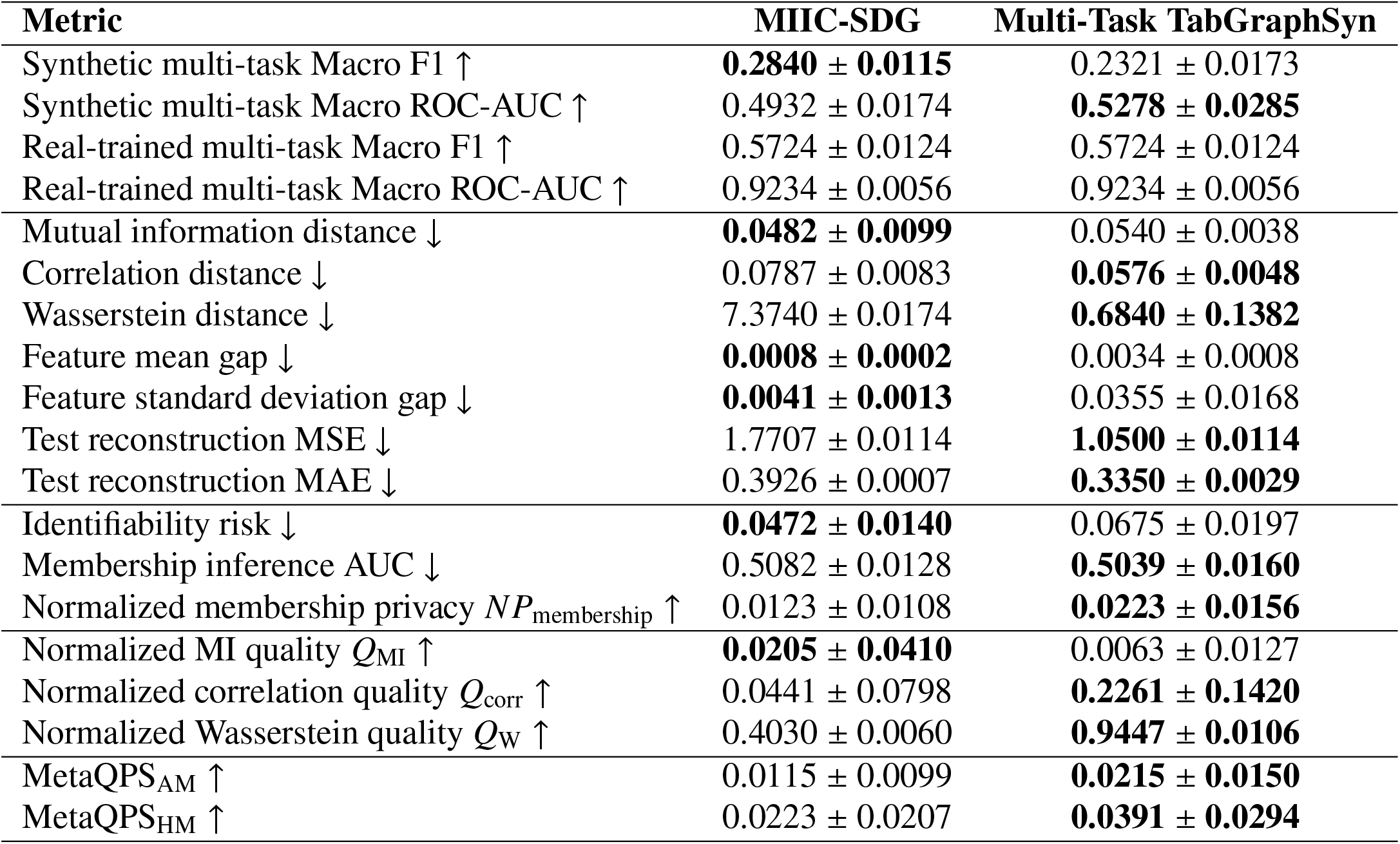
Headline comparison between MIIC-SDG and Multi-Task TabGraphSyn. Best values are shown in bold. For utility, normalized quality, normalized privacy, QPS, and MetaQPS, higher values are better. For distance- and risk-based metrics, lower values are better. Results are reported as mean ± standard deviation across four random seeds.

### 4.1. Overall Performance Trends

The most prominent result is that Multi-Task TabGraphSyn achieved the stronger overall profile across the headline metrics. Although MIIC-SDG remained competitive and, in some cases, superior on selected measures, TabGraphSyn obtained the better average rank across the full comparison set and outperformed MIIC-SDG on most of the normalized quality, privacy, and composite trade-off metrics. Most notably, TabGraphSyn achieved higher MetaQPS scores, with *MetaQPS*_*AM*_ *=* 0.0215± 0.0150 compared with 0.0115 ± 0.0099 for MIIC-SDG, and *MetaQPS*_*AM*_ *-* 0.0391 ±0.0294 compared with 0.0223 ±0.0207. These results indicate that, when utility and privacy are evaluated jointly rather than in isolation, the proposed graph-based framework provides a more favorable balance.

This distinction is important in the clinical synthetic data setting. A synthetic data generator is rarely useful if it excels on only one axis, such as fidelity or privacy, while failing on the others. The results suggest that TabGraphSyn offers a more balanced operating point, which is especially valuable for privacy-sensitive healthcare applications where both downstream usability and protection against disclosure risk must be considered simultaneously.

### 4.2. Multi-Task Utility: Divergence Between F1 and ROC-AUC

A more nuanced picture emerges when examining downstream multi-task predictive utility. MIIC-SDG outperformed TabGraphSyn in synthetic multi-task macro F1, achieving 0.2840±0.0115 compared with 0.2321 ±0.0173. However, TabGraphSyn achieved higher synthetic multi-task macro ROC-AUC, with 0.5278 ± 0.0285 compared with 0.4932 ± 0.0174 for MIIC-SDG. This divergence between F1 and ROC-AUC is consistent with the fact that these metrics capture different aspects of predictive behavior.

Macro F1 is threshold-dependent and sensitive to class imbalance and discrete decision boundaries, whereas ROC-AUC evaluates ranking quality over all possible thresholds. In the present setting, the higher ROC-AUC of TabGraphSyn suggests that its synthetic data preserve class-separating signal more effectively in a probabilistic sense, even if the default classifier thresholds do not translate that advantage into higher F1. By contrast, MIIC-SDG appears to preserve sharper decision boundaries for the evaluated classifier, which may explain its stronger macro F1 performance. This interpretation is reinforced by the utility gap results: TabGraphSyn showed a smaller degradation in macro ROC-AUC relative to real-trained performance, whereas MIIC-SDG showed a smaller degradation in macro F1.

The per-task analysis provides further insight. A task-level breakdown is provided in Table 2, which shows that MIIC-SDG achieved higher macro F1 across all three tasks, whereas Multi-Task TabGraphSyn achieved higher macro ROC-AUC for all three tasks. For all three targets, MIIC-SDG achieved higher synthetic-train/real-test F1, whereas TabGraphSyn achieved higher ROC-AUC for each task. For the 180-day opioid abuse outcome, TabGraphSyn reached a macro ROC-AUC of 0.5445 versus 0.4743 for MIIC-SDG. Similar advantages were observed for opioid concept group (0.5251 versus 0.5021) and opioid source concept group (0.5137 versus 0.5033). These results suggest that the proposed method better preserves ordinal or ranking information across all tasks, even though MIIC-SDG may produce feature-label structures that are more readily exploited by a linear classifier at fixed thresholds.

**Table 2:**
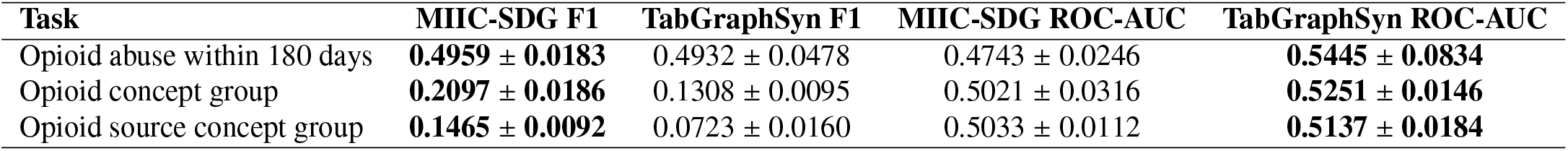
Per-task synthetic-train/real-test utility comparison between MIIC-SDG and Multi-Task TabGraphSyn. Best values for each task and metric are shown in bold.

| Task | MIIC-SDG F1 | TabGraphSyn F1 | MIIC-SDG ROC-AUC | TabGraphSyn ROC-AUC |
| --- | --- | --- | --- | --- |
| Opioid abuse within 180 days | **0.4959 ± 0.0183** | 0.4932 ± 0.0478 | 0.4743 ± 0.0246 | **0.5445 ± 0.0834** |
| Opioid concept group | **0.2097 ± 0.0186** | 0.1308 ± 0.0095 | 0.5021 ± 0.0316 | **0.5251 ± 0.0146** |
| Opioid source concept group | **0.1465 ± 0.0092** | 0.0723 ± 0.0160 | 0.5033 ± 0.0112 | **0.5137 ± 0.0184** |

From a clinical machine learning perspective, this distinction matters. In many deployment scenarios, particularly in risk stratification, ranking quality is often as important as hard classification performance because thresholds may later be tuned for different use cases. Therefore, the higher ROC-AUC of TabGraphSyn strengthens the case that it captures more transferable multi-task signal from the original EHR data.

### 4.3. Distributional Fidelity and Structural Preservation

The fidelity results indicate that TabGraphSyn more effectively preserved several aspects of the multivariate feature distribution. The strongest advantage was observed for Wasserstein distance, where TabGraphSyn achieved 0.6840 ± 0.1382 compared with 7.3740 ± 0.0174 for MIIC-SDG. This is a substantial improvement and suggests that the synthetic data generated by TabGraphSyn are much closer to the real data in the low-dimensional global geometry captured by principal component projections. TabGraphSyn also outperformed MIIC-SDG on correlation distance (0.0576 ± 0.0048 versus 0.0787 ± 0.0083), indicating better preservation of pairwise dependence structure.

These gains were reflected in the normalized quality metrics as well. TabGraphSyn achieved markedly higher *Q*_Wasserstein_(0.9447 ± 0.0106 versus 0.4030 ± 0.0060) and higher *Q*_corr_ (0.2261 ± 0.1420 versus 0.0441 ± 0.0798). These results are consistent with the intended role of the patient similarity graph and graph-aware embeddings in the proposed architecture. By learning patient-level relational structure before generative modeling, TabGraphSyn appears better able to preserve cohort-level organization and nonlinear patterns that are not captured by feature-level dependency reconstruction alone.

However, MIIC-SDG showed stronger performance on mutual information distance (0.0482 ± 0.0099 versus 0.0540 ± 0.0038) and normalized MI quality (0.0205 ± 0.0410 versus 0.0063 ±0.0127). This finding is not surprising given the design of MIIC-SDG, which explicitly models conditional dependencies among variables through an information-theoretic graphical framework. In other words, MIIC-SDG appears better at preserving certain discrete dependency relationships, whereas TabGraphSyn better preserves global geometry and correlation structure.

A similar pattern is seen in the feature-level gap metrics. MIIC-SDG achieved smaller feature mean and standard deviation gaps, indicating closer alignment of first-order marginals. By contrast, TabGraphSyn performed better on reconstruction MSE and MAE, which suggests that its learned latent representation retains stronger local similarity to real samples in the standardized feature space. Taken together, these results suggest a trade-off in the type of fidelity each method preserves: MIIC-SDG more closely matches marginal feature summaries and mutual information structure, whereas TabGraphSyn better captures broader multivariate geometry and graph-informed relational organization.

### 4.4. Privacy Risk and Normalized Privacy Scores

The privacy findings are similarly mixed but informative. MIIC-SDG achieved lower identifiability risk than TabGraphSyn (0.0472 ± 0.0140 versus 0.0675 ± 0.0197), indicating that real patients were less likely to have extremely close synthetic counterparts under the identifiability criterion. This suggests that MIIC-SDG may be somewhat more conservative in terms of patient-level closeness to the original dataset.

In contrast, TabGraphSyn achieved slightly lower membership inference AUC (0.5039 ± 0.0160 versus 0.5082 ± 0.0128), implying marginally stronger resistance to attacks that attempt to determine whether a real sample belonged to the training set. Although the difference is modest, it becomes more meaningful in the normalized privacy scores, where TabGraphSyn achieved a higher *NP*_membership_ (0.0223 ± 0.0156 versus 0.0123 ± 0.0108). For normalized identifiability, both methods effectively collapsed to zero, indicating that neither approach meaningfully improved over the random baseline on that specific normalized criterion.

These results suggest that the two models expose different privacy profiles rather than one uniformly dominating the other. MIIC-SDG appears somewhat stronger on direct nearest-neighbor identifiability, whereas TabGraphSyn offers a slight advantage against membership inference. This is consistent with the broader theme of the study: graph-based generative modeling may better preserve usable signal and global structure, but doing so can sometimes increase local resemblance to real samples. At the same time, the privacy results show that this increased fidelity does not translate into uniformly worse privacy exposure, particularly when evaluated through membership inference.

### 4.5. Quality-Privacy Trade-off

The strongest evidence in favor of the proposed method lies in the trade-off metrics. TabGraphSyn outperformed MIIC-SDG on several pairwise QPS metrics, particularly those combining Wasserstein- or correlation-based quality with membership privacy. For example, TabGraphSyn achieved higher QPS_Wasserste,Mem_ (0.0431 ± 0.0301 versus 0.0234 ± 0.0203) and higher QPSC_Corr,Mem_ (0.0390 ± 0.0284 versus 0.0095 ± 0.0130). It also achieved a non-zero QPS_MI, Mem_, whereas MIIC-SDG remained effectively zero on that metric.

These results are important because they show that the superiority of TabGraphSyn is not driven by utility alone or by privacy alone, but by the combined balance between the two. In practice, this is the criterion most relevant for synthetic healthcare data sharing. A method that slightly underperforms on one isolated utility metric may still be preferable if it preserves more clinically useful structure overall while maintaining comparable privacy risk. The higher MetaQPS values of TabGraphSyn support exactly this interpretation.

### 4.6. Why Graph-Based Multi-Task Generation May Help

The performance profile of TabGraphSyn is consistent with the design of the model. The patient similarity graph allows the model to incorporate neighborhood information and cohort-level organization before entering the generative stage. This is especially relevant in EHR data, where risk patterns often arise through shared comorbidity structure, treatment histories, and utilization trajectories rather than through independent feature effects. The graph convolutional encoder captures this relational context, and the conditional variational autoencoder then generates synthetic samples in a latent space informed by both individual attributes and graph structure.

This design likely contributes to the observed gains in correlation preservation, Wasserstein distance, and downstream ROC-AUC. Moreover, because the framework was explicitly adapted to a multi-task setting, it is better aligned with preserving signal relevant to multiple opioid-related targets simultaneously rather than a single prediction endpoint. Even though labels were appended through bootstrap sampling rather than jointly decoded, the synthetic features still appear to preserve a richer ranking structure across the three tasks than the MIIC baseline.

### 4.7. Robustness Across Seeds

The results across four random seeds indicate that the overall conclusions are not driven by a single favorable run. Both methods showed some variability, particularly in normalized and composite metrics, but the broad pattern remained stable: MIIC-SDG tended to lead on F1, feature moment gaps, and mutual information distance, whereas TabGraphSyn consistently led on ROC-AUC, Wasserstein-based quality, correlation-based quality, and the aggregate MetaQPS measures. This stability strengthens the interpretation that the observed differences reflect real methodological characteristics rather than noise due to stochastic optimization alone.

### 4.8. Implications for Privacy-Preserving OUD Research

From an applied perspective, these findings suggest that Multi-Task TabGraphSyn is a promising approach for synthetic data generation in opioid use disorder research, especially when the goal is to preserve downstream modeling utility across multiple related outcomes while maintaining a reasonable privacy profile. In a sensitive clinical domain such as OUD, synthetic data may support benchmarking, model development, or collaborative analysis when direct data sharing is not feasible. The present results indicate that graph-based multi-task synthesis can improve the overall quality-privacy balance beyond what is achieved by an information-theoretic Bayesian network approach alone.

### 4.9. Limitations

Several limitations should be acknowledged. First, the evaluation was conducted on a single opioid cohort derived from the *All of Us* dataset, and the observed patterns may not generalize unchanged to other clinical domains or data-generating processes. Second, although the proposed method was designed for a multi-task setting, the target variables were incorporated through bootstrap target sampling rather than joint conditional decoding, which may have limited task-specific F1 performance. Third, the baseline comparison focused exclusively on MIIC-SDG. While this choice was intentional and scientifically justified, broader benchmarking against additional modern synthetic tabular generators would help position the method more comprehensively. Finally, the privacy analysis relied on empirical attack metrics rather than formal privacy guarantees such as differential privacy.

### 4.10. Future Directions

Future work should investigate direct joint label-aware generation within the decoder so that multi-task label structure is modeled end-to-end rather than appended after generation. It would also be valuable to extend the framework to richer temporal or event-sequence representations, evaluate it across multiple external clinical datasets, and examine whether stronger privacy-preserving training strategies can improve identifiability without sacrificing utility. More broadly, the results motivate further exploration of graph-based generative modeling as a practical framework for privacy-aware synthetic clinical data sharing.

## Data Availability

All data produced in the present study are available upon reasonable request to the authors

https://databrowser.researchallofus.org/

